# Student nurses’ views on shift patterns: what do they prefer and why? Results from a Tweetchat

**DOI:** 10.1101/2021.07.09.21260255

**Authors:** Chiara Dall’Ora, Jessica Sainsbury, Chris Allen

## Abstract

**Aim:** To understand student nurses’ views around shift patterns.

**Design:** Qualitative study

**Method:** We held a Tweetchat in May 2019, where we asked questions around the frequency of 12-hour shifts working on placement; schedule flexibility while on placement; which shift patterns they preferred and why. Data from the Tweetchat were analysed to identify emerging themes and inductively develop a coding frame.

**Results:** Seventy-three nursing students participated in the Tweetchat. The majority (68%) of respondents reported that they work 12-hour shifts on placements. We identified three main overarching themes: “Achieving a personal equilibrium”; “Meeting the needs of the care environment”, “Factors affecting negotiation capacity”. Data highlighted a conflict for most students, where they preferred 12-hour shifts because of more time off for study, paid work, and leisure, while acknowledging 12-hour shifts had a negative effect on fatigue, exhaustion, and their ability to achieve self-care (i.e. poor diet, no exercise).

## DESIGN

This was a qualitative study of student nurses’ views on shift patterns, obtained through a TweetChat.

## METHOD

We held a Tweetchat on 2 May 2019, hosted by *The Student Nurse Project* Twitter account. Twitter is increasingly used in healthcare research, not only as a means of dissemination of studies’ findings, but also to collect data on a public platform, where is offers the opportunity to gather a large amount of data in a fast and cost-free manner (Sinnenberg et al., 2017) The views of student nurses have also previously been gathered successfully for research, using such an approach.(Clyne, Pezaro, Deeny, & Kneafsey, 2018; Richardson, Grose, Nelmes, Parra, & Linares, 2016)

The Tweetchat occurred over an hour, and was advertised by The Student Nurse Project for a week before the Tweetchat. A pre-chat blog was written, where all potential participants were informed that the data from the Tweetchat would be used to inform a piece of research.

The questions, which were sent 12 minutes apart, can be seen in **Table 1**. Participants were able to respond to questions for a further month, at which point the data were extracted. This also allowed participants to delete their responses, if they later decided they did not want to be included in the research.

**Table 1.** Questions asked during Tweetchat

| Question Number | Question |
| --- | --- |
| 1 | As a student nurse, how often do you work 12-hour shifts on placement? |
| 2 | As a student nurse, do you have a say in your rota while on placement? To what extent do you have a say? |
| 3 | As a student nurse, what shifts and shift patterns do you prefer and why? |
| 4 | As a student nurse, focusing on 12-hour shifts, what do you like about this type of shift? |
| 5 | As a student nurse, remaining focused on 12-hour shifts, what do you dislike about this type of shift? |

## ANALYSIS

Data from the Tweetchat were analysed using inductive thematic analysis (Braun & Clarke, 2006) to identify emerging themes and develop a coding frame. All data were subjected to this coding process. Data were analysed by all authors, who met frequently to discuss the emerging themes. We also extracted quantitative data, including the proportion of student nurses working long shifts, the proportion of students who can self-schedule, and which shift patterns they prefer.

## ETHICS

We obtained ethics approval for this study from the XXX Ethics Committee (Approval ID: 48131). Participants’ confidentiality could not be maintained during the Tweetchat, as Twitter is a publicly available platform that anyone can access. Participants were aware that their tweets would be used for research and therefore, whilst consent was not formally taken, it was implied through their participation. We did not retain any personal data of participants when extracting, analysing, and reporting data.

## INTRODUCTION

The debate on whether nurses should work 12-hour shifts has been ongoing for more than 40 years (No author, 1975). Large multi country studies of thousands of nurses across Europe and the US have univocally shown negative impacts of 12-hour shifts on both patient care and on outcomes of nurse wellbeing and performance (Clendon & Gibbons, 2015; Dall’Ora, Ball, et al., 2019; Dall’Ora, Griffiths, Ball, Simon, & Aiken, 2015; Dall’Ora, Griffiths, et al., 2019; Griffiths et al., 2014; Stimpfel & Aiken, 2013; Stimpfel, Lake, Barton, Gorman, & Aiken, 2013; Stimpfel, Sloane, & Aiken, 2012). Nonetheless, anecdotal reports that many nurses prefer working long shifts are frequent, with better work-life balance and costs often cited as main motivators (Ball, Dall’Ora, & Griffiths, 2015). Qualitative and mixed-methods studies on the views of nursing staff on shift patterns have started to emerge in recent years, and they offer a range of views. A study of 25 healthcare assistants found that many preferred long shifts, but only when some work factors were in place, including adequate staffing levels, breaks during the shift and supportive team climate (Thomson, Schneider, & Hare Duke, 2017). Forty-two nurses reported preferring 12-hour shifts because of better continuity of care, enhanced communication with patients, and, ultimately, patient outcomes. A further beneficial aspect of 12-hour shifts were reduced childcare and commute costs, and nurses concluded that the higher fatigue they experienced during long shifts was due to high patient acuity (Haller, Quatrara, Letzkus, & Keim-Malpass, 2018).

## BACKGROUND

Despite growing qualitative evidence around nurses’ views of 12-hour shifts, research on student nurses’ perceptions and opinions of shift patterns is currently missing. In the UK, student nurses tend to exhibit different characteristics from staff nurses, especially students who enrolled after the NHS Bursary Scheme, which covered University tuition fees of £9,000 per year and offered students financial support, was abolished in 2018. Student nurses face multiple challenges, including combining academic study and placement demands. In particular, students with families and caring responsibilities are confronted with competing demands in managing and balancing their student and family life (O’Brien, Graham, & O’Sullivan, 2017). It is not uncommon for student nurses to seek part-time employment, with reports of up to 35 hours per week (Rochford, Connolly, & Drennan, 2009) to support their living expenses and studies (Hasson, McKenna, & Keeney, 2013).

The extent to which any of these factors influence student nurses’ preferences and views on shift patterns is unknown. Furthermore, it is unclear as to the extent to which student nurses are involved in long shift patterns, and the amount of flexibility they have in negotiating their shift pattern. This study aims to explore which shift patterns student nurses in the UK work, which factors are important for student nurses to design their schedule, where allowed, and what their views of different shift patterns are.

## RESULTS

Seventy-three nursing students took part in the Tweetchat, and generated 252 Tweet interactions. Sixty-eight percent of respondents reported that they mostly work 12-hour shifts on placements, while 27% identify a mixture of long and short shifts, often depending on the type of placement (e.g. longer shifts in acute ward placements as opposed to short shifts in community placements). Only two students reported working mostly 8-hour shifts on placement. For data on the degree of flexibility and ability to influence their placement rota and shift patterns preferences, please see **Table 2**.

**Table 2.**
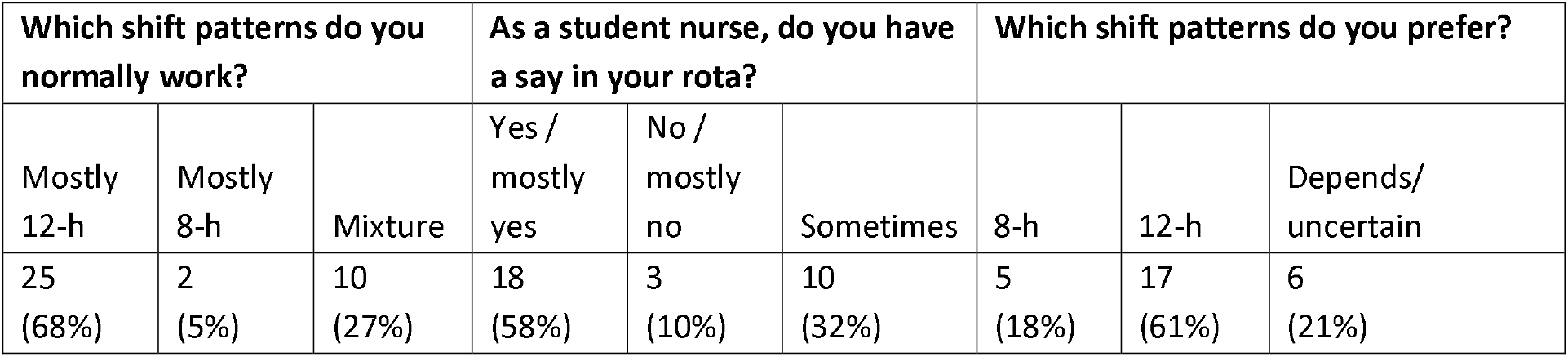
Student nurses report of typical shift patterns, degree of say when planning rotas, and shift preferences

| Which shift patterns do you normally work? |  |  | As a student nurse, do you have a say in your rota? |  |  | Which shift patterns do you prefer? |  |  |
| --- | --- | --- | --- | --- | --- | --- | --- | --- |
| Mostly 12-h | Mostly 8-h | Mixture | Yes / mostly yes | No / mostly no | Sometimes | 8-h | 12-h | Depends/ uncertain |
| 25 (68%) | 2 (5%) | 10 (27%) | 18 (58%) | 3 (10%) | 10 (32%) | 5 (18%) | 17 (61%) | 6 (21%) |

Three major themes emerged when looking at students’ narratives: achieving a personal equilibrium; care environment needs; factors affecting negotiation capacity.

### Achieving a personal equilibrium

When expressing their preference for 8 or 12-hour shifts, it became apparent that for students achieving a personal equilibrium was of vital importance. The need for personal equilibrium was expressed in terms of financial, learning, personal network work, and physical and mental health needs. 12-hour shifts both helped and hindered the attainment of these.

#### Financial needs

A large number of students agreed that when finances were considered, 12-hour shifts were preferable in terms of reduced travel costs (for example, fuel and parking to cover 3-4 days per week) and increased capacity to do paid work (working longer shifts gives more ‘whole days off’):

> *“(I prefer) long days, 3 days a week-gives me time to get a shift in for £. And cost is massive for me-I struggled last year doing ⅘days a week financially for petrol/parking (STN9)”*

Some students mentioned that besides travel time and ability to engage in more extra paid work, finding parking was also an issue on 8-hour shifts, especially when these were later in the day. Childcare costs were also relevant for some students:

> *“I like more days off to study or work, less costs on travel and parking and less childcare costs (STN13)”*

#### Learning needs

When considering learning needs, especially practice learning, students appeared to prefer 12-hour shifts, presumably due to the longer consecutive time they offered on a shift, where they could engage in less fragmented activities, compared to 8-hour shifts:

> *“I think (12-hour shift working) it has taught me the routine of the ward well, I have also been able to form relationships with patients as you can spend more time with them (STN43)”*

Due to having more days off with 12-hour shifts, students reported preferring them in terms of having more time to complete university work:

> *“I prefer 12-hour shifts. They give me more days off to complete academic work (STN3)”*

For some students, however, working 12-hour shifts was so tiring that university work felt compromised:

> *“If I do three (12-h shifts) in a row the tiredness can then affect my ability to do uni work (STN13)”*

The participants’ views on 12-hour shifts, reflect some of the challenges experienced by students enrolled on healthcare programmes, with regards to balancing the practice related requirements of the course, alongside the programme’s academic requirements. Currently students need to complete at least 2,300 practice hours across the length of their programme. This is complemented with a further 2,300 theory hours. Because of these demands, it is common for nursing degrees to run into university holiday periods, where students on other degrees have opportunities to explore extracurricular activities.

#### Personal network work

Having time to engage in personal networks, including family, caring, social, and “life admin” activities was a relevant factor in students’ apparent preference for 12-hour shifts.

> *“(With) 12 hours, (I have) more quality time with my family on my days off. (STN12)”*

However, some acknowledged that traditional childcare arrangements were not ideal when working 12-hour shifts:

> *“I prefer longer days, allowing me more time off to spend with my kids, (although finding childcare for shift working is difficult) (STN21)”*

For some, working long shifts, meant “not see[ing] [their] kids for days”, and not being able to engage in activities with them during the day:

> *“I don’t see the kids for days. 7am-8pm, asleep when I go to work and asleep when I get home. Not easy (STN40)”*
>
> *“Mother guilt, I feel guilty leaving before (my children) wake, guilty that it’s nearly their bedtime when I get home and being tired on my days off (STN21)”*

It was common for students to face role conflict. This was most visible when students discussed shifts preventing them from doing the school run and other activities that were aligned to the role of parent. Whilst such conflicts have been noted in the context of nursing elsewhere (Yildirim & Aycan, 2008; Zandian, Sharghi, & Moghadam, 2020), there is currently limited recognition of such conflicts in pre-registration nursing programmes. Such conflicts often resulted in a preference towards longer shifts. However, when considering activities such as housework and personal appointments, students indicated a preference for 8-hour shifts:

> *“I prefer 8 hours. Yes, it takes up 5 days of my week but it’s a week pattern I’m used to. If I’m on earlies, it means I finish at 3*.*15 and still have time to do other things like book appointments after 3 (STN18)”*

#### Physical and Mental Health

Whilst many student nurses reported preferring 12-hour shifts due to the reduced travel costs and having more days off (often to allow them to work elsewhere), they mostly acknowledged that working 12-hour shifts (especially where these were used to allow for remunerated employment) paid quite a severe toll on their physical and mental health:

> *“I dislike 12-hour nights. I’m so exhausted a week after. On days: you still likely get 1 break which you would on an 8 and so you’re hungry and can’t think straight which leads to mistakes. 12 hours + commute is a long time, not enough time to sleep (STN46)”*
>
> *“I’ll do three in a row I sometimes feel I lose touch with my ’real life’-the tiredness can then affect my ability to do uni work (STN13)”*
>
> *“I much prefer the 5 days personally. I like part of my day to not be about sleep and work. I love nursing but I want more than that in my day. It would be nice to have the option to do the shorter days (STN69)”*

The literature links both shift work, and long working hours, (both commonly experienced by the included students) with negative outcomes, including burnout and fatigue, the genesis of chronic illness, obesity, and reduced job performance (Caruso, 2014; Dall’Ora et al, 2015). Besides fatigue, and sleeping patterns students voiced their concerns about how long shifts impact their ability to follow a healthy diet and get enough physical exercise:

> *“I really struggle with eating on long days, I always plan to take a meal and snacks. But I end up eating 4 meals or just a pile of rubbish. I have gained nearly a stone and a half since I started University! (STN43)”*
>
> *“I also find it harder to schedule in regular and consistent exercise week-on-week (STN22)”*

These concerns are echoed elsewhere in the literature and it is known that shift work, more generally, is associated with negative health outcomes, including metabolic syndrome and obesity (Zhang et al., 2020).

Impact on mental health was recognised by many, who recognised how working long shifts and, in addition, booking in extra paid work, and assignments, could lead to feelings of exhaustion and eventually burnout:

> *“12*.*5 shifts also not healthy as a student nurse when most of us also have paid work. I have worked 60+ hrs when on placement. It’s a recipe for exhaustion, fatigue and ultimately burnout. No time for meals or sleep (STN45)”*

Such trends reflected the large number of unpaid placement hours, the cost of attending university, and the need to engage in supplementary paid employment to meet students’ financial needs and continue the programme. The financial challenges associated with undergraduate nursing programmes are noted across the literature (Cleary, Horsfall, Baines, & Happell, 2012; He, Turnbull, Kirshbaum, Phillips, & Klainin-Yobas, 2018; Van Hoek, Portzky, & Franck, 2019) and were very visible in the students narratives.

### Meeting needs of the care environment

When students responded to questions regarding pros and cons of 12-hour shifts, they also reflected on how these shift patterns promote or hinder meeting the needs of the care environment, in terms of continuity of patient care and workload demands.

#### Patient continuity and holistic care

As seen in literature with registered nurses (Baillie & Thomas, 2019), several students displayed a preference for 12-hour shifts in terms of continuity of care. This was expressed mostly around the concept of “getting to know the patients better”, completing more care and avoiding miscommunication of information due to fewer handovers:

> *“Personally, I prefer 12*.*5 hour shifts as it gives me a chance to get to know my patients for the day and follow their treatment throughout the day to the full extent, and it saves multiple transfers of care which could cause miscommunication (STN26)”*

Some students thought that 12-hour shifts also helped provide holistic care to patients.

> *“I like how I can spend more time with service users, understand and empathise with them more which ensures I can provide holistic, person centred care as I know their goals, values etc. (STN3)”*

A few students raised the objection that while care continuity during the day might be increased, the higher number of days off resulting from 12-hour shifts meant that students did not see patients for four days, which impacted on continuity of care:

> *“It also means 4 days I’m not seeing people in my care and less stability for them (STN57)”*

#### Workload

Students were conscious that when it came to the impact of different shift patterns, high workload and lack of breaks were a determining factor for disliking 12-hour shifts:

> *“It can be physically exhausting working for 12*.*5 hours straight, especially on busy understaffed shifts when there’s not much time for a break. Although supernumerary, I still feel the need to help out as if a substantive sometimes. It’s not the staff’s fault it’s busy! (STN45)”*

However, some student nurses felt that given the high pressures of their placements, they were compelled to help as much as possible, both because of concerns for how the team saw them, as well as concerns for their colleagues in practice and concerns for patients, meant they felt compelled to work long shift patterns:

> *“It can be hard to be supernumerary when you are still being judged for how well you behave as a nurse on a ward. There is so much pressure to get things done, and keep up with every other member of staff on the ward (STN63)”*
>
> *“Sometimes I wonder how my mentor copes when I am not there, often it’s just been the 2 of us on a bay and we’re both rushing around. Today I went for student lecture and felt I was abandoning her, she was trying to do admissions, discharges, switch beds and obs (STN64)”*

### Factors affecting negotiation capacity

Our question around ability to choose and negotiate shift patterns elicited a variety of responses in student nurses’ ranging from no flexibility to complete ability to design their rotas provided NMC requirements for total hours were met. This raises issues of equity, between nursing students, on the same programme, but also across different programmes, as it highlights the role of chance, in determining how likely a student is able to negotiate shifts to accommodate meeting personal and occupational needs. We offer some quotes to illuminate the aspects that influence negotiation capacity.

#### Capacity to negotiate

Some students seemed to be given advice by their universities regarding shift patterns. Whilst no students reported mandatory shift lengths, 12-hour shifts were discouraged by some higher education institutions (HEIs).

> *“We are recommended to do 2 x7*.*5 hr shifts a week but on ward placement I would do 12 hr (STN30)”*

It was unclear how this related to student’s individual capacity to negotiate shifts. However, informal more flexible arrangements were available to some students. Some students were able to negotiate shifts around part time work and other commitments; though this was not a universal experience, raising issues of equitability in this aspect of many nursing programmes. Some students described how different placements had different stances regarding shift flexibility:

> *“Depends on the placements, some get quite annoyed if you want to change only one day whereas the one I’m on now lets you change them around whenever as they say its our responsibility to get the hours done we need and to make sure we work with our mentors (STN6)”*
>
> *“In theory we work the shifts given to us, but in my own experience all my shift-based placements have been very accommodating if I wanted to switch a few shifts around (STN22)”*

This research was carried out during the standards, which required students to spend at least 40% of their practice hours, working alongside their mentor, which influenced the shifts the students were able to do:

> *“I’ve had to work when my mentor is working but normally this has been fine and I can pick which shifts of theirs suit me (STN3)”*

Whilst this still allowed some negotiation, particularly if the mentor worked fulltime, those whose mentors were part time, or where there were lots of students placed in the same setting, invariably ended up having to follow the unsocial working patterns of their mentors:

> *“I’ve been able to have quite a lot of flexibility where I’ve had full time mentors, where they’ve been part time I’ve had to basically do every shift they’re on so I’ve ended up with tonnes of nights (STN46)”*

Whilst this requirement limited some negotiation capacity, particularly where the students’ mentor was part time, participants often prioritised working alongside their mentor as much as possible to get the best learning opportunities. This tended to lead to students working 12-hour shifts, especially in acute environments where such shifts are now normalised (Dall’Ora, Ball, et al., 2019).

Indeed, 12-hour shifts now appear to be something of a hidden curriculum (Raso et al., 2019) in many practice settings.

> *“I try to follow my mentor’s shifts and he has had all 12*.*5 hours, but I do get to choose my shifts, I just choose to stick with him (STN26)”*

In addition, a few students described how asking for changes in shift patterns had to be approached with careful and considered diplomacy:

> *“Our rotas are made for us and most wards will let you change a few shifts if need be-providing you go about it the right way and have a reasonable excuse for changing (STN42)”*
>
> *“All my placements have been very flexible with my shifts. You just have to approach them in the right way and remember flexibility works two ways (STN12)”*

In some cases having personal commitments could be used as leverage to change shift patterns:

> *“I’ve always had a say in my rota whilst on placement-however this may be due to me having caring responsibilities that sometimes require flexibility. I’ve never had a negative experience when asking to change shifts or when I can only work certain days (STN24)”*
>
> *“Generally I’ll work what I’m given the only request changes which impact my family such as sports day, parents’ evenings, health appointments or award ceremonies (STN44)”*

However, such a strategy was not always successful, and reflected a tension between the requirements of the course, and the need to maintain other roles:

> *“I’ve never had a placement take my kids into account with off duty. I have missed plays and concerts. And always be told to get used to it (STN62)”*

Overall, it appears that the most determining aspect of students’ flexibility is the ward/clinical setting where students are in placement and the normalised shift pattern of that environment. However, other factors may impact on students’ ability to negotiate their shift patterns.

#### Limited capacity to negotiate

We further explored which aspects were detrimental to student ability to negotiate shift patterns. Students acknowledged how having placements in settings with fixed service hours was a limiting factors:

> *“Every ward based placement was 3x 12*.*25 hour shifts (day and night shift). All community placements would be 8 hour shifts and you’d do these over 5 days (STN16)”*
>
> *“Monday to Friday placements such as out patients there’s no negotiation (STN44)”*

Some placements did not allow students to work long shifts, hence students experienced no negotiation capacity:

> *“ 12 hours on a ward if I can, but I’ve had one ward that didn’t let students do 12-hour days and one that didn’t do 12-hour days because although it was classed as a ward it was research so did short days (STN23)”*

Compared to employed colleagues, student nurses have less say in where they work, and how they work. Registered nurses, acting as employees, are better able to negotiate specific patterns of work, that consider personal needs (I.e. childcare and caring responsibilities) and/or financial reasons (such as enhanced antisocial pay). The ability to negotiate patterns of work does not appear to be available for student nurses in the same way.

## DISCUSSION

This study was the first to elicit student nurses’ views and experiences around shift patterns on placement. By analysing thematically 252 tweets by 73 student nurses, we found that the majority work and prefer long shifts, and most can influence their rota on placement-though this is not consistently experienced, raising issues of equity. When exploring what students like and dislike about 12-hour shifts, two main priorities were uncovered – namely *achieving a personal equilibrium and meeting the needs of the care environment*.

When it came to work-life balance, students appeared to prefer long shifts, as these were offering more days off which could be used for engaging in social activities, caring for children and dependants, and save money on parking and travel. Our findings mirror previous qualitative evidence from registered nurses and nursing assistants, where one of the reasons for preferring 12-hour shifts was the extra days off they offer (Baillie & Thomas, 2019; Haller et al., 2018).

Notwithstanding the preference for long shifts, several students reported feeling tired when working these rotas. Participants often described themselves as “exhausted”, “drained”, “fatigued” when working 12-hour shifts. Burnout and fatigue are also more common among registered nurses and healthcare assistants who work long shifts (Barker & Nussbaum, 2011; Dall’Ora et al., 2015; Suter et al., 2020; Thomson et al., 2017), indicating that the potential of long shifts to lead to negative wellbeing outcomes for student nurses is high. Recovery after 12-hour shifts is challenging, with nurses describing the first few days off as merely recuperation time, where they strive to make up for the lost sleep and the cumulative fatigue (Chen, Davis, Daraiseh, Pan, & Davis, 2014; Gifkins, Johnston, Loudoun, & Troth, 2020; Han, Trinkoff, & Geiger-Brown, 2014; Linsey M. Steege, Pasupathy, & Drake, 2014). However, a factor that makes working long shifts possibly more demanding for students is that they need to also complete academic work, including studying, and writing assignments, and for many students, complete additional paid shifts. This is of potential concern, especially considering that some students in the Tweetchat reported working more than 60 hours a week, which is considered to be an excessively long working week (Harma et al., 2015) and enhances the risk of lack of recovery from long shifts, which is essential to avoid the development of chronic fatigue (Chen et al., 2014; L. M. Steege, Pasupathy, & Drake, 2018). Such excessive hours breach European working time directive (Directive 2003/88/EC, 2003), but are often necessary for students to be able to complete their programmes.

Despite expressing feelings of exhaustion, most students did not seem to believe the quality of care they provided was somehow affected when working long shifts; it appeared that student nurses were aware of the fatigue *after* the shift, but only a few acknowledged the *within* shift fatigue caused by 12-hour shifts. This neglect of in-shift fatigue resembles that outlined by the *“Supernurse culture”* construct, whereby nurses report experiencing fatigue, but believe it is a standard component of nursing; therefore, nurses feel they have the tools and skills to operate despite being fatigued, similar to superheroes having special powers (L. M. Steege & Rainbow, 2017). Only a minority of students acknowledged that working more than 8-hour shifts meant they could not practice to their best standards. Moreover, the *Supernurse culture* was also highlighted when students mentioned feeling compelled to help the understaffed and under resourced wards by working long shift patterns. While the desire to relieve pressure from strained health services is commendable, it appeared to motivate students to prioritise the placements’ needs over their fundamental and learning needs. Recent pressures, brought about by the COVID-19 pandemic, have further tested the appropriateness of the decision to remove the bursary, with many final year students halting their academic study, to join the front-line response to the pandemic, as non-supernumerary, paid members of the healthcare workforce. This highlights the difference in the way nurses receive education, compared to other degree fields.

Far from identifying 12-hour shifts as a potential predictor for poor quality of care, most students felt that working long shifts increased continuity of care and holistic care, since they would be seeing the same patients for longer on the same day. Some students shared that having only one handover instead of two, across the 24-hours of the day led to a lower risk of miscommunication between shifts. The anecdotal argument of increased continuity of care seems to disappear when looking at surveys of >30,000 nurses across 12 countries, where it was found that 12-hour shifts were not associated with increased continuity of care, and that no important information was missed at handover when shorter shifts were worked (Dall’Ora et al., 2020). In addition, a few students reported that having more days off could also lead to reduced continuity of care, highlighting how no shift patterns are ideal to maintain continuity of care (Sparbel & Anderson, 2000).

When focusing on capacity to negotiate shift patterns, most students reported being able to influence their schedule; among the factors which might have increased capacity to negotiate there were personal commitments including childcare and part time work, although these were not always perceived by placements as valid reasons to have flexible shift patterns, hence there was always an element of chance.

Students’ accounts elicited the existence of *a “placement lottery”*, according to which a student may get completely opposite stances regarding work hours depending on where they are on placement. While this lottery construct does not apply to placements with fixed service hours, where choice of shift patterns is by definition constrained, it was seen in several students’ recounts of feeling powerless in some placements, and being involved in designing their rotas in other placements. Higher work time control can be beneficial for nurses, in terms of their sickness absence (Turunen et al., 2020) and wellbeing (Nijp, Beckers, Geurts, Tucker, & Kompier, 2012), although any positive effects appear to be negated when healthcare professionals opt to work long shifts (Karhula et al., 2020; Tucker, Bejerot, Kecklund, Aronsson, & Akerstedt, 2015). Nonetheless, there seems to be inequity in the degree of choice that students’ nurses are given when it comes to shift patterns. Providing more flexibility may open up nursing as a career choice to those with caring responsibilities.

### Limitations

Compared to in-depth interviews, deploying a Tweetchat does not allow substantial breadth and depth. Nonetheless, this study was the first to elicit student nurses’ views on shift patterns, and thus can serve as a starting point for future studies seeking to clarify the lived experiences, needs, and preferences of student nurses. We did not collect any demographic data. Future research should aim to collect such information to ensure the sample is representative of different universities and geographical areas within the UK. Whilst it was intended that all participants were currently student nurses, this could not be guaranteed.

## CONCLUSIONS

The views and preferences around student nurses’ shift patterns are varied, but most students prefer working long shifts due to the resulting increased days off, used mainly to study and write assignments, and, importantly, to achieve paid work. While students acknowledge that the toll of long shifts on their physical and mental health is high, they seem to prioritise the extra days off over their wellbeing. This highlights the increased pressures students are subject to, whereby they do not only need to achieve a work-life balance, but a work-university-life balance, sometimes whilst being in practice for more than 60 hours a week. The little awareness of the impact of fatigue on wellbeing, performance and patient safety is worrying, and should be addressed in nursing programmes from a theory perspective, but also when placements are planned. The inequality of students’ choice and negotiation around shift patterns should also be addressed by nursing programmes, who might act as advocates of equal student choice. Ideally, given the high risk of harm that fatigued students might expose themselves and their patients to, any efforts both at a national and university level to financially aid students should be encouraged and pursued.

## Data Availability

No data are available

## Acknowledgments

We thank all the student nurses who spent their free time participating in the Tweetchat and shared their opinions with us.

